# High amino-acid intake in early life is associated with systolic but not diastolic arterial hypertension at 5-years of age in children born very preterm

**DOI:** 10.1101/2023.08.11.23294001

**Authors:** Jean-Christophe Rozé, Justine Bacchetta, Alexandre Lapillonne, Farid Boudred, Jean-Charles Picaud, Laetitia Marchand-Martin, Alexandra Bruel-Tessoulin, Jérome Haramba, Valérie Biran, Dominique Darmaun, Pierre-Yves Ancel

**Affiliations:** Department of Neonatal Medicine, Nantes University Hospital, Nantes, France; Reference Centre for Rare Kidney Diseases, INSERM 1033 Research Unit, Hospices Civils de Lyon, Lyon 1 University, France; Department of Neonatal Medicine, Assistance Publique Hopitaux de Paris, Necker Enfants Malades Hospital, Paris, France; Department of Neonatology, Faculté de Médecine, Aix-Marseille Université, Marseille, France; Department of Neonatology, Hospices Civils de Lyon, Lyon, France and Laboratoire CarMen, INSERM, INRA, Université Claude Bernard Lyon1, Pierre-Bénite, F69310, France; Université Paris Cité, Sorbonne Paris-Nord, Inserm, INRAE, CRESS, Obstetrical Perinatal and Pediatric Epidemiology Research Team, EPOPé, F-75006 Paris, France; Pediatric Nephrology Unit, Department of Pediatrics, Nantes University Hospital, Nantes, France; Pediatric Nephrology Unit, Department of Pediatrics, Bordeaux University Hospital, Bordeaux, France; Neonatal Intensive Care Unit, Assistance Publique-Hôpitaux de Paris, Robert Debré Children’s Hospital, Paris, France; UMR 1280, INRAE-Nantes Université, Nantes, France

## Abstract

**BACKGROUND:** The life course of individuals born very premature is a topic of increasing concern as their neonatal survival has dramatically increased. In a national, prospective, population-based birth cohort, EPIPAGE-2, we observed that amino-acid intakes greater than 3.5 g/kg/day at day 7 after birth were independently associated with higher intelligence quotient at 5 years. As the association between high early amino-acid intake and later HBP in this population is debated, we assessed blood pressure (BP) at 5 years.

**METHODS:** This cohort was initiated in 2011, and approved by ethics committees. Eligible infants were those born between 24 and 29 weeks of gestation and alive on day7 after birth. Infants were distributed in two groups of 717 infants matched on propensity score on whether or not they were exposed to high amino-acid intake (>3.5 g/kg/d at Day7); 455 control term infants were also enrolled. Assessment at 5-year occurred from September 2016 to December 2017. A value ≥ 95^th^ percentile of reference values for age and height defined systolic and/or diastolic HBP.

**RESULTS:** BP at five years of age was assessed for 389 and 385 children in exposed and non-exposed groups. Rates (%) of systolic HBP were 18.0% (95%CI, 14.5 to 22.2), 13.3% (95%CI, 10.3 to 17.0), and 8.5% (95%CI, 6.5 to 11.1) in exposed, non-exposed preterm infants, and term infants, respectively; and 9.0% (95%CI, 6.6 to 12.3), 10.2% (95%CI, 7.5 to 13.6), 5.4% (95%CI, 3.8 to 7.6) % for diastolic HBP, in exposed, non-exposed and term-born groups, respectively. Perinatal characteristics of exposed and non-exposed infants were similar, except for nutrition intake at day3 and day7 after birth. Exposure to high early amino-acid intake, and maximal serum creatinine between day3 and day7 were two independent risk factors for systolic HBP (aOR, 1.60 [95% CI, 1.05 to 2.43] and aOR, 1.59 [95% CI, 1.12 to 2.26] by 50 µmol/L, respectively) but not for diastolic HBP (aOR, 0.84 [95% CI, 0.50 to 1.39] and aOR, 1.09 [95% CI, 0.71 to 1.67] by 50 µmol/L, respectively).

**CONCLUSIONS:** These observations confirm the risk for HBP in young children born very preterm. Higher amino-acid intake and creatinine values in the first week of life were associated with childhood systolic HBP. These results suggest that mechanisms to childhood systolic HBP involves neonatal renal challenge by high amino-acid intake or dysfunction.

## INTRODUCTION

The life course of individuals born very premature has become a topic of increasing importance as their survival beyond neonatal period has dramatically increased over the last few decades. The risk for developing systemic arterial high blood pressure (HBP) is increased in this population, but the underlying mechanism(s) are poorly understood. ^1,2,3^

Epidemiologic evidence suggests early increased amino-acid intake could be associated with later HBP in general population. This association could be mediated by altered kidney function.^4^ Literature suggests postnatal nutritional intakes could be determinants of later high blood pressure at 6.5 years of age in infants born very preterm as well.^5^ Concerns have been raised that not only low intakes but also high intakes in infancy might contribute to higher blood pressure and increased cardiovascular risk later in life in this high-risk population.^6^

In a birth national prospective population-based cohort, (*Etude Epidémiologique sur Petits Ages Gestationnels* [EPIPAGE-2]), we observed that amino acid intake, above 3.5g/kg/day at day 7 after birth, was independently associated with higher intelligence quotient at 5 years in a propensity score matched cohort.^7^ As the association between high early amino-acid intake and later HBP is debated in this population, we assessed BP at 5-year in this matched cohort.

## METHODOLOGY

### Study Population

Very preterm were enrolled in EPIPAGE 2 cohort. Recruitment took place at birth in all NICUs in France from April 1st to December 31th, 2011. Eligible children were those born between 24+0 and 29+6 weeks+days of gestation, admitted to the NICU, alive at 7 days after birth, and with information available regarding amino acid intake at 7 days after birth. Baseline characteristics at birth and during the first week, as well as the maximum creatinine value obtained between day 3 and day 7 after birth. were prospectively recorded during neonatal hospitalization. Children were followed from September 1, 2016, to December 31, 2017. The study was approved by the Nationa Data Protection Authority, the Consultative Committee on the Treatment of Information on Personal Health Data for Research Purposes, and the Committee for the Protection of People Participating in Biomedical Research.^8^

Term born children were from the ELFE (*Étude Longitudinale Française depuis l’Enfance*) cohort, followed with the EPIPAGE-2 follow-up protocol. The ELFE cohort is a contemporary French cohort of more than 18000 children born in 2011, in 344 randomly selected public and private maternity units in metropolitan France.^9^ Recruitment and data collection occurred only after families had received information and provided oral informed consent to participate in the study. This study followed the Strengthening the Reporting of Observational Studies in Epidemiology (STROBE) reporting guideline for cohort studies.

### Amino Acid Intake at 7 Days After Birth

Preterm infants were separated into 2 groups, exposed and nonexposed, based on whether they had been prescribed a high amino acid intake (defined as 3.51-4.50 g/kg/d at 7 days after birth)), as recommended in 2011 on physiological basis,^10^ by the American National Institute of Child Health and Human Development,^11^ the European Society of Paediatric Gastroenterology, Hepatology and Nutrition.^12^ In the EPIPAGE-2 study, information about nutritional intake was recorded at days 3, 7, and 28 and at hospital discharge. All data were prospectively collected during NICU hospitalization.

### Outcomes

The primary outcomes were systolic and diastolic HBP defined as a value > 95th percentile of reference values for age and height.^13^ The secondary outcomes were systolic and diastolic BP considered as continuous variables. Standard operating procedures were used for resting BP measurements.^14,15^ Blood pressure (mmHg) was measured noninvasively during an outpatient visit at 5 years of age by automatic oscillometer with a blood pressure monitor or with a manual sphygmo-manometers. Children were seated in a quiet examination room for at least 5 minutes before BP assessement. All BPs were assessed by trained, certified staff and done in agreement with Fourth Task Force recommendations of technique and cuff sizes.^14^ Three consecutive BP measurements were performed 2 minutes apart. The reported value was the mean of the 2 closer values. If only 2 measurements were performed, the reported value was the mean of these two values. In case of only one measurement, this value was reported. Z-score of weight at 5-year were based on WHO curves.

### Statistical Analysis

The main analysis included 1789 children born preterm with complete data. We analyzed the association between the exposure and the primary outcome using a propensity score approach^16^ to control for observed confounding factors that might have consequences for both group assignment (exposed vs nonexposed) and outcome. The propensity score of each infant was defined as the probability of having an amino acid intake greater than 3.50 g/kg/d based on the infant’s individual observed covariates. The score was estimated using a logistic regression model, with amino acid intake greater than 3.50 g/kg/d at day 7 as the dependent variable with regard to baseline maternal, infant, and NICU characteristics. Birth weight z-scores were based on Olsen curves. The proportion of participants with missing data ranged from 0% to 8.5%, exceeding 4.0% only for data on Apgar score at 5minutes and maternal educational level. Missing data were treated as a separate category. The primary analysis was based on propensity score matching. We used a 1:1 matching algorithm without replacement to match exposed and nonexposed newborns based on propensity score withina caliper of 0.2 SD of the logit of the propensity score.^17^ Imbalance after matching was checked (Table 1). First, we compared rate of systolic and diastolic HBP between exposed and non-exposed preterm infant groups, between exposed preterm infant group and control born term group and between non-exposed preterm infant group and control term-born group. Second, we calculated Odds ratios (ORs) to quantify the association between high amino acid intake at day 7, maximum serum creatinine between day 3 and day 7 and primary outcomes (systolic and diastolic HBP) using logistic regression analysis fit by generalized estimating equations to account for paired data.^18^

**Table 1.** Baseline characteristics.

|  | No./Total ( ) or Mean (SD) |  |  |  |  |  |  |
| --- | --- | --- | --- | --- | --- | --- | --- |
|  | Overall Cohort (N = 1789) |  |  |  | Matched Cohort (N = 1434) |  |  |
| Characteristics | Nonexposed<br>N =851<br>n (%) | Exposed<br>N=938<br>n (%) | Standardized<br>difference |  | Nonexposed<br>N=717<br>n (%) | Exposed<br>N=717<br>n (%) | Standardized<br>difference |
| **Maternal level of education** |  |  |  |  |  |  |  |
| Information missing | 137 (16.10) | 103 (10.98) | 15.01 |  | 95 (13.25) | 95 (13.25) | 0 |
| less than high school diploma | 241 (28.32) | 231 (24.63) | 8.37 |  | 194 (27.06) | 193 (26.92) | 0.32 |
| High school diploma | 157 (18.45) | 175 (18.66) | 0.54 |  | 137 (19.11) | 134 (18.69) | 1.07 |
| Higher than high school diploma | 316 (37.13) | 429 (45.74) | 17.55 |  | 291 (40.59) | 295 (41.14) | 1.12 |
| **Gestational age in weeks.** mean (SD) | 27.16 (1.55) | 27.18 (1.46) | 1.33 |  | 27.21 (1.55) | 27.17 (1.47) | 2.65 |
| **Birthweight z-score.** mean (SD)* | -0.10 (0.99) | -0.09 (1.01) | 1.00 |  | -0.07 (±0.97) | -0.07 (±1.01) | 0.00 |
| **Sex** |  |  |  |  |  |  |  |
| Male | 469 (55.11) | 460 (49.04) | 12.11 |  | 386 (53.84) | 376 (52.44) | 2.81 |
| Female | 382 (44.89) | 478 (50.96) |  |  | 331 (46.16) | 341 (47.56) |  |
| **Leading causes of preterm delivery** |  |  |  |  |  |  |  |
| Information missing | 20 (2.4) | 36 (3.8) | 8.61 |  | 17 (2.37) | 19 (2.65) | 1.79 |
| Twins or triplets | 288 (33.84) | 287 (30.60) | 6.94 |  | 242 (33.75) | 225 (31.38) | 5.06 |
| Singleton with preterm labor | 239 (28.08) | 251 (26.76) | 2.96 |  | 195 (27.2) | 207 (28.87) | 3.72 |
| Singleton with preterm rupture of membranes | 127 (14.92) | 164 (17.48) | 6.95 |  | 113 (15.76) | 114 (15.90) | 0.38 |
| Singleton with vascular disorders and FGR | 66 (7.76) | 71 (7.57) | 0.71 |  | 55 (7.67) | 56 (7.81) | 0.52 |
| Singleton with vascular disorders. without FGR | 76 (8.93) | 82 (8.74) | 0.67 |  | 63 (9.00) | 63 (9.03) | 0.00 |
| Singleton with placental abruption | 11 (1.29) | 13 (1.32) | 0.87 |  | 10 (1.43) | 10 (1.43) | 0.00 |
| Singleton with isolated fetal growth retardation | 24 (2.82) | 34 (3.62) | 4.53 |  | 22 (3.14) | 23 (3.30) | 0.80 |
| **Antenatal corticosteroids** |  |  |  |  |  |  |  |
| Information missing | 30 (3.53) | 33 (3.52) | 0.05 |  | 30 (4.18) | 29 (4.04) | 0.71 |
| No | 143 (16.80) | 160 (17.06) | 0.69 |  | 114 (15.90) | 122 (17.02) | 3.02 |
| Incomplete cure | 163 (19.15) | 130 (13.86) | 14.27 |  | 116 (16.18) | 115 (16.04) | 0.38 |
| Yes | 515 (60.52) | 615 (65.57) | 10.48 |  | 457 (63.73) | 451 (62.90) | 1.72 |
| **Caesarean section** |  |  |  |  |  |  |  |
| Information missing | 13 (1.53) | 8 (0.85) | 6.27 |  | 5 (0.70) | 6 (0.84) | 1.60 |
| Yes | 530 (62.28) | 563 (60.02) | 4.64 |  | 450 (62.76) | 435 (60.67) | 4.30 |
| **APGAR $\geq 7$ at 5 minutes after birth** |  |  |  |  |  |  |  |
| Information missing | 77(9.05) | 54 (5.76) | 12.59 |  | 52 (7.25) | 50 (6.97) | 1.09 |
| Yes | 586 (68.86) | 684 (72.92) | 8.95 |  | 505 (70.43) | 507 (70.71) | 0.61 |
| **Regular intestinal transit during the first week after birth** |  |  |  |  |  |  |  |
| Information missing | 34(4.00) | 43(4.58) | 2.86 |  | 31(4.32) | 31(4.32) | 0 |
| Yes | 430 (50.53) | 499 (53.20) | 5.35 |  | 359 (50.07) | 379 (52.86) | 5.58 |
| **Acute kidney failure** |  |  |  |  |  |  |  |
| Information missing | 23 (2.70) | 31 (3.30) | 3.52 |  | 20 (3.18) | 19 (2.75) | 2.54 |
| Yes | 107 (12.57) | 59 (6.29) | 17.31 |  | 59 (8.23) | 56 (7.81) | 1.55 |
| **Surfactant** |  |  |  |  |  |  |  |
| • Information missing | 4 (0.47) | 2 (0.21) | 4.47 |  | 2 (0.28) | 2 (0.28) | 0 |
| • No | 143 (16.80) | 149 (15.88) | 2.49 |  | 127 (17.71) | 117 (16.32) | 3.70 |
| • 1 dose | 493 (57.93) | 578 (61.62) | 7.53 |  | 421 (58.72) | 430 (59.97) | 2.55 |
| • >2 doses | 211 (24.79) | 209 (22.28) | 5.9 |  | 167 (23.29) | 168 (23.43) | 0.33 |
| **Assisted ventilation at day 7** |  |  |  |  |  |  |  |
| Information missing | 3 (0.35) | 11 (1.17) | 9.45 |  | 2 (0.28) | 4 (0.56) | 4.33 |
| Yes | 409 (48.06) | 366 (39.02) | 18.31 |  | 314 (43.79) | 282 (39.33) | 9.06 |
| **Patient volume of NICU where the infant was hospitalized at day 7** |  |  |  |  |  |  |  |
| • <20 infants | 183 (21.50) | 113 (12.05) | 25.50 |  | 124 (17.29) | 108 (15.06) | 6.06 |
| • 21-30 infants | 150 (17.63) | 227 (24.20) | 16.21 |  | 137 (19.11) | 145 (20.22) | 2.79 |
| • 31-40 infants | 132 (15.51) | 111 (11.83) | 10.73 |  | 102 (14.23) | 104 (14.50) | 0.77 |
| • >40 infants | 386 (45.36) | 487 (51.92) | 13.15 |  | 354 (49.37) | 360 (50.21) | 1.68 |

\* birth weight z-score based on Olsen's curves <sup>[40]</sup>

Sensitivity analyses were performed among the overall cohort, after adjustment for gestational age, sex, birth weight *z* score, and adjusting for risk factor for HBP, using the inverse probability treatment weighting method,^19^ and among the matched cohort but restricted to blood pressure measurements performed with an automatic blood pressure monitor.

Complementary analyses were performed using systolic and diastolic BPs as continuous variables. First, we measured the Spearman correlation between systolic and diastolic BPs and macronutrient intake as a continuous variable in the matched cohort. In the overall cohort, we used general linear equations and adjustment for gestational age, sex, and birth weight *z* score, weighted by the inverse of the propensity score. A pathway analysis was performed to find out whether the relationship between high early amino-acid intake and later BP is mediated by the 5-year weight Z-score.

All tests were 2-sided, and p< .05 were considered statistically significant. All analyses were performed using SPSS®, version 26. Data were analyzed from January 15th to May 15th, 2023.

## RESULTS

### Study population

Among the 2136 preterm infants born between 24+0 and 29+6 gestation admitted to NICUs during the study period, 170 died within the first 7 days, leaving 1966 alive at day 7. Among these, information about amino-acid intake at day 7 after birth was available in 1789 infants. Among the 1789 infants, 938 received more than 3.5g/kg/day (exposed group) and 851 did not (non-exposed group) (Figure 1). These infants were hospitalized in 63 NICUs, where the percentage of exposure varied from 0–100% (eFigure 1 in the Supplement). Differences between the exposed and non-exposed infants are presented in Table 1.

**Figure 1.**
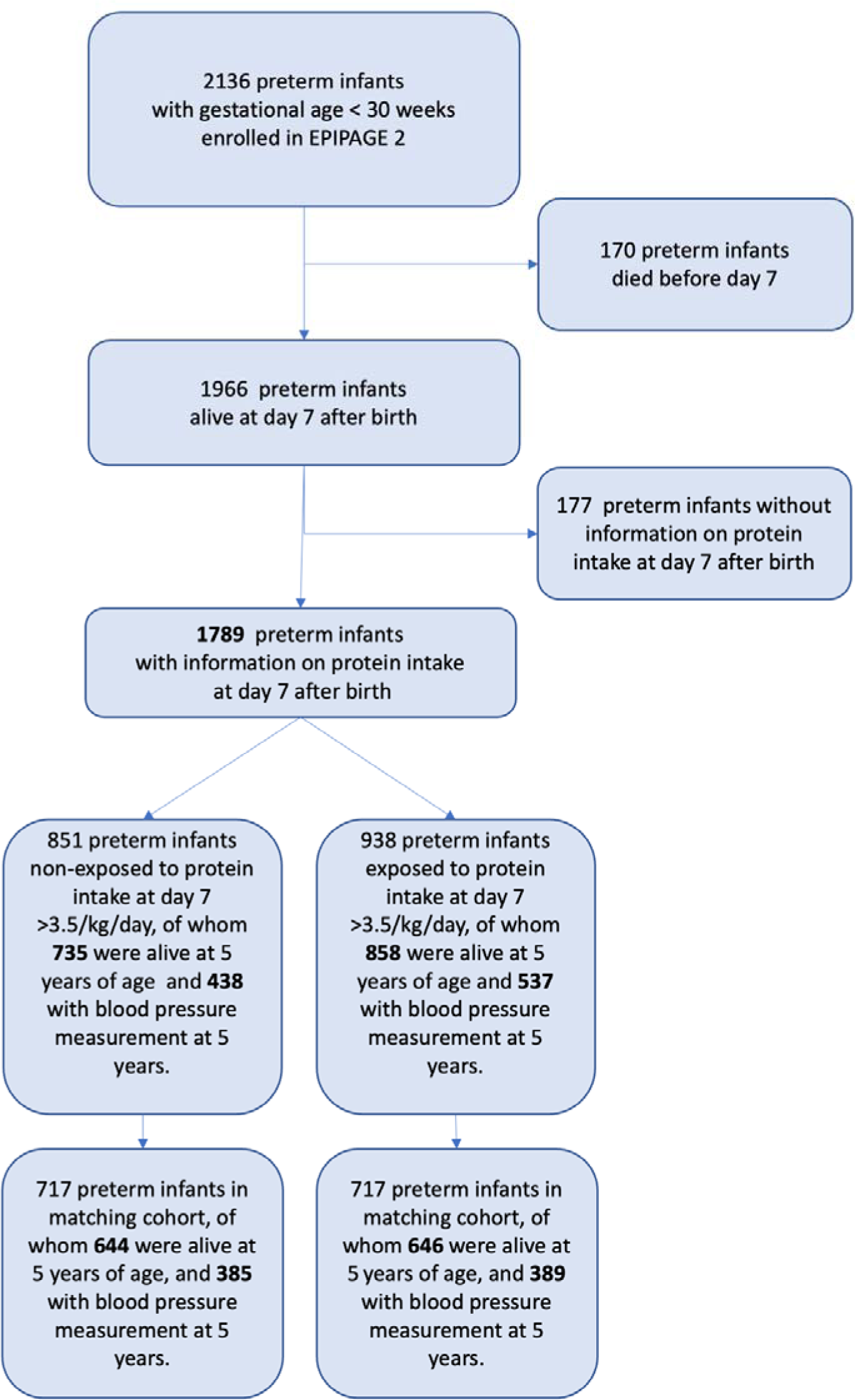
Flow chart.

### Propensity matched analysis

Propensity scores were calculated in 1789 very preterm infants. Distributions of propensity scores are summarized in eFigure 2 in the Supplement. Propensity scores ranged from 0.095 to 0.922. The area under the receiver operating characteristic curve for the propensity score model was 0.67 (95% confidence interval [CI], 0.64 to 0.69), eFigure 2. Among the 1789 preterm infants, 1434 could be matched, with 717 in each group, exposed and non-exposed. The matched groups were found to be well-balanced for all recorded baseline variables (Table 1). Maximal creatinine observed in plasma between day 3 and day 7 after birth, was significantly lower in exposed group (Table 2). Characteristics of nutrition intake at day 3, 7 and 28 after birth and outcome at 36 weeks of postmenstrual age are indicated in Table 2. At 36 weeks of postconceptional age and at discharge, nutrition intake were similar.

**Table 2.**
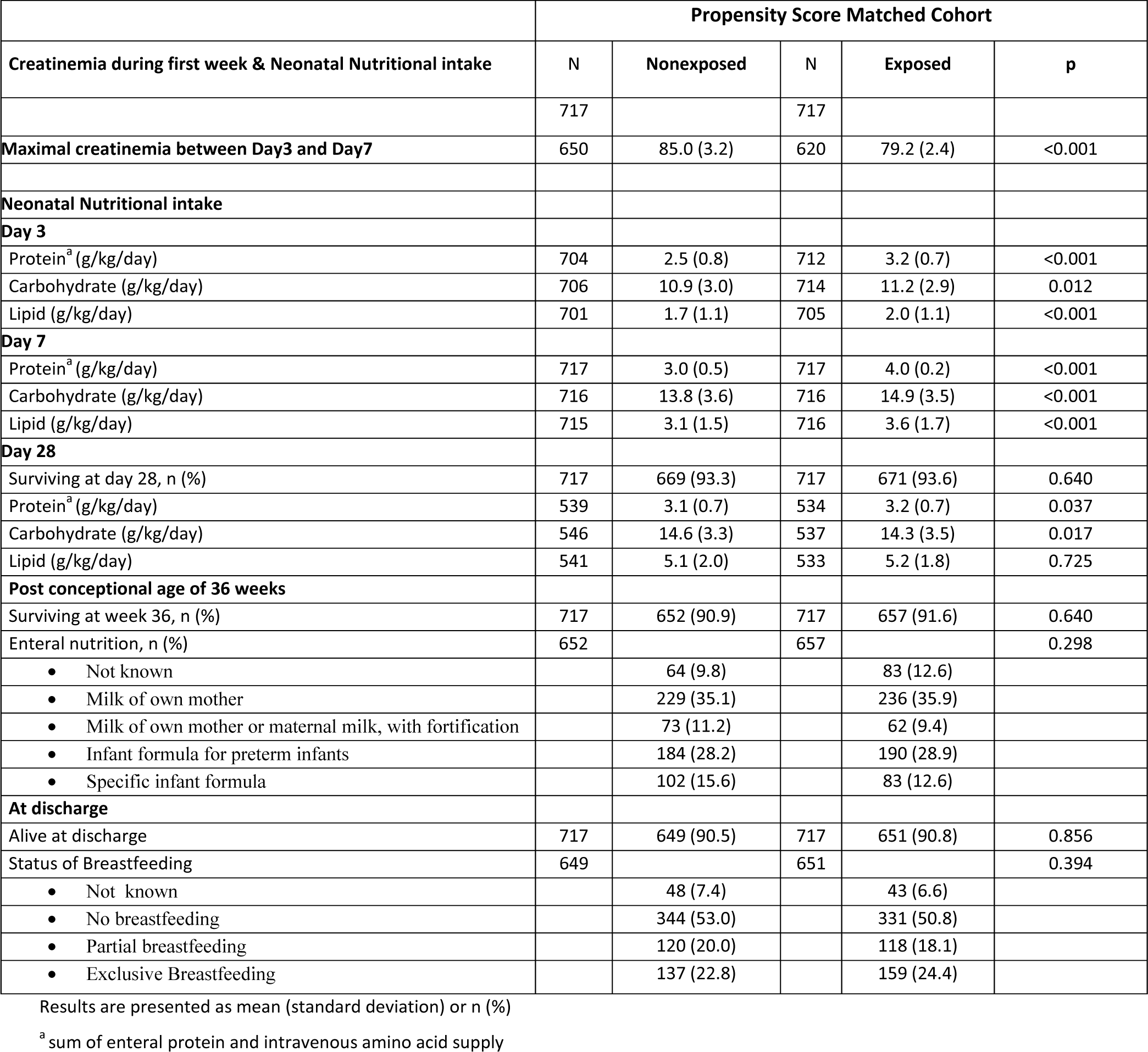
Neonatal Nutritional intake in the Non-Exposed and Exposed groups.

In the matched cohort, at five years, rates of children alive were similar in the 2 groups. The number of measurements of blood pressure was 3 for 354 and 358, 2 for 21 and 19, and only 1 for 10 and 12 children in the non-exposed and exposed groups, respectively. The mode of measurement was automatic oscillometric method in 291/385 (75.6%) and 308/389 (79.2%) of Non-exposed and Exposed groups, respectively, p=0.023. The mean systolic blood pressure was higher in the Exposed group: 100.2 (10.6) versus 98.6 (10.4) mmHg, p=0.036; but not the mean diastolic blood pressure: 59.1 (8.9) versus 58.7 (8.6) mmHg, p=0.615 (Table 3). Systolic HBP tended to be more frequent in Exposed group, 18.0% (70/388) versus 13.3% (51/384), p=0.069 but not diatolic HBP rate, 9.0% (35/388)) versus 10.2% (39/384), p=0.592. In control group born at term, rates of systolic and diastolic HBP were 8.5% (49/574) and 5.4% (31/574), respectively. Thus, compared with control group, the rate systolic HBP was greater in the 2 preterm groups, whether exposed (18.0 % versus 8.5%, p<0.001) or not exposed (13.3% versus 8.5%, p=0.019). Similarly, diastolic HBP rate was more common (9.0% and 10.2% in exposed and non-exposed preterm groups, respectively), compared with 5.4% (p=0.03 and P=0.006)) in controls.

**Table 3.** Growth characteristics and blood pressure at 5 years in the Non-Exposed and Exposed groups.

| Growth characteristics and blood pressure at 5 years | Propensity Score Matched Cohort |  |  |  |  |
| --- | --- | --- | --- | --- | --- |
|  | N | Nonexposed | N | Exposed | p |
|  | 717 |  | 717 |  |  |
| **At Birth** |  |  |  |  |  |
| Gestational age (weeks) | 717 | 27.1 (1.5) | 717 | 27.1 (1.5) | 0.599 |
| Birth weight, (g) | 717 | 1000 (250) | 717 | 990 (250) | 0.501 |
| Birth weight Z-score |  | -0.07(0.00) |  | -0.07 (0.10) | 0.993 |
| Length, (cm) | 663 | 35.4 (3.0) | 680 | 35.6 (3.0) | 0.566 |
| Birth length Z-score |  | -0.15 (0.97) |  | -0.09 (0.97) | 0.274 |
| Body mass index (BMI), | 663 | 7.9 (1.2) | 680 | 7.8 (1.1) | 0.123 |
| Birth BMI Z-score |  | 0.07 (1.13) |  | -0.01 (1.04) | 0.186 |
| **At Discharge** |  |  |  |  |  |
| Post menstrual age (weeks) | 643 | 39.0 (4.5) | 646 | 38.8 (4.0) | 0.340 |
| Weight, (g) | 634 | 2730 (580) | 638 | 2800 (600) | 0.053 |
| Discharge weight Z-score |  | -1.27 (0.84) |  | -1.13 (0.86) | **0.003** |
| Length, (cm) | 612 | 45.7 (3.2) | 609 | 45.9 (3.2) | 0.165 |
| Discharge length Z-score |  | -1.92 (1.03) |  | -1.77 (1.09) | **0.019** |
| Body mass index (BMI), | 612 | 13.0 (1.48) | 609 | 13.1 (1.54) | 0.154 |
| Discharge BMI Z-score |  | -0.04 (0.9) |  | 0.05 (0.9) | 0.131 |
| Delta weight Z-score between birth and discharge | 634 | -1.23 (0.83) | 638 | -1.08 (0.82) | **<0.001** |
| Delta length Z-score between birth and discharge | 557 | -1.78 (0.99) | 566 | -1.70 (1.02) | 0.176 |
| Delta BMI Z-score between birth and discharge | 557 | -0.21 (1.25) | 566 | 0.02 (1.20) | **0.003** |
| **At 5 years** |  |  |  |  |  |
| Weight, (g) | 397 | 18.5 (3.1) | 408 | 19.1 (3.6) | **0.008** |
| Weight Z-score |  | -0.30 (1.13) |  | -0.07 (1.27) | **0.008** |
| Height, (m) | 394 | 1.11 (0.05) | 407 | 1.12 (0.06) | 0.140 |
| Height Z-score |  | -0.12 (1.11) |  | 0.03 (1.21) | 0.067 |
| Body mass index (BMI), | 394 | 14.9 (1.6) | 407 | 15.3 (2.0) | **0.011** |
| Birth BMI Z-score |  | -0.27 (1.12) |  | -0.12 (1.25) | 0.072 |
| Delta weight Z-score between discharge and 5 years | 397 | 1.03 (1.16) | 397 | 1.17 (1.26) | 0.107 |
| Delta height Z-score between discharge and 5 years | 394 | 1.90 (1.20) | 394 | 1.89 (1.89) | 0.842 |
| Delta BMI Z-score between discharge and 5 years | 394 | -0.18 (1.27) | 394 | 0.30 (1.46) | 0.051 |
| **All blood pressure measurements** |  |  |  |  |  |
| • Systolic arterial pressure, mean (SD) | 385 | 98.6 (10.4) | 389 | 100.2 (10.6) | **0.036** |
| • Systolic high blood pressure for sex and height, n (%) | 384 | 51 (13.3) | 388 | 70 (18.0) | 0.069 |
| • Diastolic arterial pressure, mean (SD) | 385 | 58.7 (8.6) | 389 | 59.1 (8.9) | 0.615 |
| • Diastolic high blood pressure for sex and height, n (%) | 384 | 39 (10.2) | 388 | 35 (9.0) | 0.592 |
| **Blood pressure measurements by automated oscillometric methods** | 291 |  | 308 |  |  |
| • Systolic arterial pressure, mean (SD) | 291 | 99.3 (10.2) | 308 | 101.4 (10.2) | **0.013** |
| • Systolic high blood pressure for sex and height, n (%) | 290 | 39 (13.4) | 307 | 63 (20.5) | **0.022** |
| • Diastolic arterial pressure, mean (SD) | 291 | 59.0 (8.1) | 308 | 59.1 (8.6) | 0.801 |
| • Diastolic high blood pressure for sex and height, n (%) | 290 | 24 (8.3) | 307 | 275(8.8) | 0.953 |

Results are presented as mean (standard deviation) or n (%)

Among preterm infants, exposure to a high amino acid intake at day 7 after birth, and maximal creatinine observed in plasma between day 3 and day 7 after birth, were significantly associated with systolic HBP (adjusted Odds Ratio [aOR], 1.60; 95% CI, 1.05 to 2.43 and aOR=1.59; 95%CI 1.12 to 2.26, respectively), but not with diastolic HBP, (aOR =0.84; 95% CI, 0.50 to 1.39 and aOR=1.09; 95%CI 0.71 to 1.67, respectively) (Figure 2, part A).

**Figure 2.**
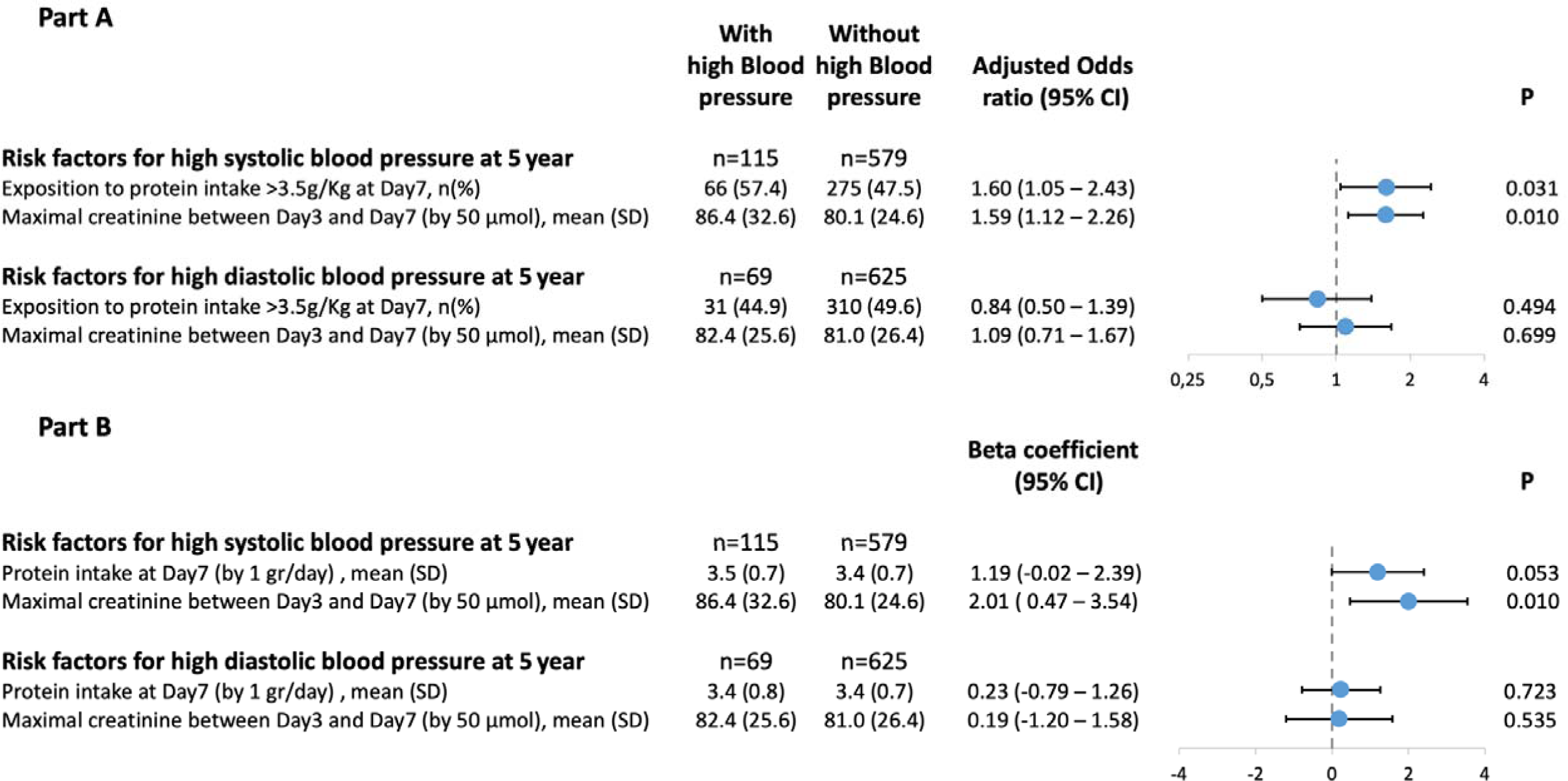
Relationship between blood pressure at five years and 2 risk factors: amino-acids intake at day 7 after birth and maximal creatinine observed between Day3 and Day 7 after birth, in matched cohort. Part A: Multivariable analysis of surviving with high blood pressure. Part B: Beta coefficient between blood pressure and the 2 risk factors

In addition, we observed a nearly significant or a significant linear relationship between amino acid intake at day 7 after birth or maximal creatinine observed in plasma between day 3 and day 7, and systolic BP as continuous variable: adjusted beta =1.00 mmHg per 1 gr /kg of amino acid intake; 95%CI −0.19 to 2.71 and adjusted beta =2.01 mmHg per 50 µmol of creatinine; 95%CI 0.42 to 3.60 respectively (Figure 2, part B). But, we did not observe any significant correlation between amino acid intake at day 7 or maximal creatinine observed in plasma between day 3 and day 7, and diastolic blood pressure. The pathway analysis showed that the relationship between high early amino-acid intake and later systolic BP was not mediated by the 5-year weight Z-score (eFigure 3).

### Sensitivity analyses

These associations were consistent across sensitivity analyses performed in the overall cohort or in the matched cohort but restricted to blood pressure measurements performed with an automatic blood pressure monitor.

**• Inverse probability of treatment weighting analysis in the overall cohort**

Exposure to a high amino acid intake at day 7 after birth and the maximal plasma creatinine observed between day 3 and day 7 after birth, were significantly associated with systolic HBP (aOR= 1.51; 95% CI, 1.02 to 2.24 and aOR=1.52; 95%CI 1.08 to 2.13, respectively), but not with diastolic HBP, (aOR =0.89; 95% CI, 0.57 to 1.39 and aOR=1.02; 95%CI 0.68 to 1.53, respectively) after adjustment for gestational age, sex, birth weight Z-score and weighted by the inverse of the propensity score (eFigure 4, Part A).

**• Matched cohort restricted to automatic blood pressure measurements**

The mean systolic blood pressure was higher in the Exposed group: 101.4 (10.2) versus 99.3 (10.2) mmHg, p=0.013; but not the mean diastolic blood pressure: 59.1 (8.6) versus 59.0 (8.1) mmHg, p=0.801. Systolic HBP was more frequent in Exposed group, 20.5% (63/307) versus 13.3% (39/290), p=0.022 but not diatolic HBP rate, 9.0% (/388)) versus 10.2% (39/384), p=0.592. Exposure to a high amino acid intake at day 7 after birth and the maximal creatinine observed in plasma between day 3 and day 7 after birth, were significantly associated with systolic HBP (aOR= 1.96; 95% CI, 1.21 to 3.18 and aOR=1.78; 95%CI 1.19 to 2.66 respectively, eFigure 5, Part A), but not with diastolic HBP, (aOR =1.00; 95% CI, 0.54 to 1.85 and aOR=0.87; 95%CI 0.51 to 1.48). Moreover, the linear relationships between amino acid intake at day 7 after birth or maximal plasma creatinine between day 3 and day 7, and systolic BP (adjusted beta =1.61 mmHg per 1 gr /kg of amino acid intake; 95%CI 0.28 to 2.93 and adjusted beta 1.94 mmHg per 50 µmol of creatinine; 95%CI 0.31 to 3.57 respectively) were significant (eFigure 5, Part B). In contrast, we did not observe any significant relationship between amino acid intake at day 7 or maximal plasma creatinine between day 3 and day 7, and diastolic blood pressure. The pathway analysis showed that the relationship between high early amino-acid intake and later systolic BP was not mediated by the 5-year weight Zscore (e Figure 6).

## DISCUSSION

The findings of the current study confirm the increased risk of HBP at 5 years in a population of children born very preterm. They further reveal that the risk of systolic HBP is significantly higher in preterm infants that were exposed to higher early amino-acid intake in the first week of postnatal life. Finally, they suggest that two different mechanisms may be involved for systolic HBP and diastolic HBP, the former likely implicating the kidney since a higher “maximum plasma creatinine between day3 and day7 after birth” was a risk factor for systolic HBP, but not for diastolic HBP.

Systematic reviews and meta-analyses have reported significantly higher BP in very preterm infants, with a difference, depending on studies, ranging from 2 to 7 mmHg for systolic pressure and 2 to 3 mmHg for diastolic pressure, compared with control subjects born at full term.^1, 5, 20–27^ We here confirm the risk of higher BP at 5 years for diastolic pressure in our population of very preterm infants, regardless of amino acid intake in the first week of life. Yet the risk for systolic BP was only increased in the exposed infants. As the heights of children born very preterm are smaller once they reach late childhood, the rate of systolic HBP according to age and height is also significantly higher in non-exposed groups of children born very preterm than those born at term. Such observation is of importance as preterm-born adults have been found to have a unique left ventricular structure and function that worsens with systolic blood pressure elevation.^28^ Thus, additional primary preventive strategies specifically targeting cardiovascular risk reduction in this population may be warranted. To achieve that goal, it seems relevant to briefly discuss our understanding of the underlying pathophysiology of high systemic blood pressure.

In the current study, early exposure to higher amino-acid intake was associated with higher systolic blood pressure and increased risk of systolic HBP as a function of age and height. The association can be attributed to amino-acid intake *per se*, since baseline infant characteristics are very similar between exposed and unexposed groups in the matched cohort. Only nutritional intake at day 3 and day 7 was very different between the 2 groups, and the difference, albeit smaller, remained significant at Day 28. Though the intake of all macronutrients was higher in the exposed group, yet among the 3 macronutrients measured (amino-acid, carbohydrate and lipid), only early amino-acid intake was significantly correlated to systolic blood pressure. This is consistent with the results of the study reported by Wang et al,^29^ where a relationship between random plasma insulin levels at birth and later HBP in early childhood was observed. Though serum insulin was not measured in our study, serum insulin level likely was higher in the exposed group, since a higher intake of amino acid, particularly of branched-chain amino acids, is known to be enhance insulin secretion.^30^ In our study, higher amino-acid intake was associated only with systolic HBP, but not with diastolic HBP. Other studies that have examined the relationship between neonatal nutritional intakes and the later risk of HBP have observed either an association with diastolic HBP^31^ or no association,^30–32^ but none focused on early amino-acid intake in the first week of life.

Serum creatinine observed between Day3 and Day 7 was significantly lower in the Exposed group. At least 3 hypotheses can be raised to account for this observation. First, in theory, this could be due to an indication bias to receive higher amino-acid intake (i.e., clinicians might have been less prone to prescribe high amino acid intake in infants with a higher serum creatinemia): but this seems unlikely, given the high degree of similarity between the two matched groups (Exposed and Non-exposed). Second, a decreased rate muscle protein breakdown, resulting in a decrease in creatinine release from skeletal muscle, could have occurred in the Exposed group, since a higher amino acid intake is known to inhibit proteolysis in infants^33,34^ Third, increased glomerular filtration rate (also known as hyperfiltration) could occur in the kidneys in response to exposure to a high early amino-acid intake in the Exposed group. This latter hypothesis is consistent with animal models.^35^ As such, glomerulosclerosis would be the result of glomerular hyperfiltration on immature kidneys, which in turn could lead to secondary increased blood pressure (BP) via activation of the renin–angiotensin system as observed in rats. Moreover, as a higher maximum creatinine between day3 and day7 after birth was a risk factor for systolic HBP in our cohort, the role of kidneys is very likely, as suggested by this study performed in general population where early high protein intake was associated with later systolic hypertension and an increase in kidney volume at 11 years of age.^36^ The mechanism underlying this relationship could be the role of senescence pathways as observed in rats.^37^

Our study had several strengths and limitations. Its first strength stems from the fact that a five-years follow-up in the national population based EPIPAGE2 cohort allowed us to investigate the association between initial nutritional intake, which had been very precisely collected in a prospective fashion, and later BP at the age of 5 years. Second, prospective follow-up with 3 repeated BP measurements reduced potential bias due to inherent BP variability and measurement error. Finally, we used a propensity score approach to control as much as possible for confounding factors. Our study also had a few limitations. First, BP measurements were performed on only 60% of the children born very preterm alive at 5 years but the attrition was similar in Exposed and Non-exposed groups. Second, no blood sampling was performed and thus we have no information concerning kidney function at 5 years of age. Finally, this study remains an observational study, and, as such can only highlight associations, rather than elucidate causal relationships.

In conclusion, we confirmed the risk of HBP in mid childhood in children born very preterm and observed an association between early higher amino-acid intake and systolic HBP, whereas we had earlier reported improved cognitive outcomes at age 5 years in the same exposed group in the same matched cohort.^7^ Thus, we are still facing a persistent nutritional dilemma regarding the management of very preterm infants: how can we promote neurocognitive development and still preserve long term cardiovascular health?^38^

## Data Availability

The data are, in principle, accessible to all research teams, public, French or foreign, subject to authorisation by the cohort Data Access Committee. The new law for modernisation of the French Public Health System voted in 2016 now provides a legal framework for access to and re-use already collected cohort data by complying with ?Reference Methodology MR-004?. Therefore, only non-nominative data defined as having a low re-identification risk are accessible. Moreover, general information on research activities in the institution must be provided to the persons concerned (posting on the premises, entry in the welcome booklet, etc.) To this general information, an individual patient information must be delivered for each project in which the patient is involved or for which the patient data will be treated. As a consequence, each data access request must be submitted to the Epipage 2 Data Access Committee (DAC) that evaluates the research projects based on the following criteria: 1) methodological strengths and weaknesses (feasibility, choice of methods to achieve the objectives) 2) absence of overlap with other ongoing projects, in which case discussions with the different teams are organised 3) relevance of the requested data for the project and respect for confidentiality.

